# Integrative AI Model Combining Radiomics and Phenomics to Predict Survival in Non-Small Cell Lung Cancer Patients Treated with Immunotherapy Containing Regimen

**DOI:** 10.64898/2025.12.03.25341593

**Authors:** Junho Song, Jungmin Kim, Hangyul Lee, Ronald Min, Young Kwang Chae

## Abstract

**Purpose:** Immunotherapy has improved outcomes in non-small cell lung cancer (NSCLC), but only a subset of patients achieves durable survival benefit. Conventional biomarkers such as PD-L1 expressions have limited predictive power. This study aimed to develop an artificial intelligence (AI)–based radiomic framework integrating deep learning derived and traditional radiomic features with clinical variables to predict survival outcomes in advanced NSCLC treated with immune checkpoint inhibitor (ICI)–containing regimens.

**Methods:** Ninety-four patients with stage IV NSCLC treated with ICI-containing regimens between 2013 and 2022 were analyzed. Baseline CT scans were segmented and processed using LIFEx software, and deep radiomic features were extracted using a UNET convolutional encoder–decoder. Models were trained using nested five-fold cross-validation. Performance was evaluated using area under curve (AUC) and calibration assessment.

**Results:** The ensemble model combining clinical, traditional radiomic, and deep radiomic features achieved AUCs of 0.70 and 0.84 for 12– and 36-month overall survival (OS), and 0.65 and 0.73 for 12– and 36-month progression-free survival (PFS). For 12-month OS, the model integrating clinical variables with deep radiomic features showed the highest performance (AUC 0.78). Beyond deep features, traditional radiomic features reflecting tumor heterogeneity and morphology were the most influential predictors. Patients classified as high risk by the ensemble model had significantly shorter survival (OS HR 2.10, p = 0.005; PFS HR 1.74, p = 0.015).

**Conclusion:** Integrating deep learning–derived radiomic features with clinical data improved survival prediction in advanced NSCLC treated with ICI-containing regimens. These findings support the potential of an AI-based radiomic risk stratification model as a noninvasive tool for individualized outcome prediction, warranting prospective validation.

**Key Points:** *Question:* Can combining deep learning-derived radiomic features with clinical variables improve prediction of survival outcomes in patients with non–small cell lung cancer (NSCLC) treated with immune checkpoint inhibitor-containing regimens?

*Findings:* In a retrospective cohort of 94 patients with stage IV NSCLC, an ensemble model integrating clinical, traditional radiomic, and deep learning-derived radiomic features showed strong discrimination for 12– and 36-month overall survival with an area under curve of 0.70 and 0.84, respectively. Radiomic markers of tumor heterogeneity and morphology were among the most influential factors.

*Meaning:* Deep learning-based radiomic-clinical modeling can enhance individualized survival prediction in NSCLC.

## INTRODUCTION

Lung cancer remains the leading cause of cancer-related mortality worldwide, accounting for an estimated 1.8 million deaths annually (1). Among its subtypes, non–small cell lung cancer (NSCLC) comprises approximately 85% of all cases and presents a persistent clinical challenge due to its late presentation and limited responsiveness to conventional therapies (2). Despite ongoing advances in diagnosis and management, the five-year survival rate for lung cancer remains dismally low, typically ranging between twenty to thirty percent depending on the stage and histological subtype (3).

Immunotherapy, such as immune checkpoint inhibitor (ICIs) targeting the programmed death-ligand (PD-L1) axis, has marked a major shift in the therapeutic landscape of advanced NSCLC (4). Recently, the KEYNOTE-024 trial reported a five-year overall survival (OS) rate of 31.9% in advanced NSCLC patients with high PD-L1 expression, a result previously not reached with chemotherapy alone (5). Unlike the cytotoxic regimens, immunotherapy has shown the potential to induce durable remissions, with some patients achieving prolonged disease stability even in the metastatic setting.

Despite these promising advances, major challenges remain: the identification of patients most likely to benefit from ICI-based therapies. While PD-L1 expression remains the most widely used biomarker for immunotherapy eligibility, its predictive accuracy is imperfect. A non-negligible subset of patients with high PD-L1 expression demonstrates resistance to immunotherapy early in their course of treatment, while some patients with low or no expression of PD-L1 still derive meaningful clinical benefit (6). Alternative molecular biomarkers, such as tumor mutational burden (TMB) and microsatellite instability (MSI) status, can provide additional predictive insights but are limited by the need for tissue samples and their inability to fully reflect intratumoral heterogeneity (7; 8). Overall, it is estimated that up to 70% of patients with advanced NSCLC fail to achieve durable response to immunotherapy, calling for more accurate and comprehensive tools (9; 10).

Immune evasion in lung cancer is a complex, multiscale process involving molecular, cellular, and tissue-level mechanisms. At the tissue level, one prominent mechanism involves the formation of a tumor microenvironment that physically restricts immune cell access to malignant cells. The accumulation of organized fibroblasts in the peritumoral stroma has been shown to physically restrict CD8+ T-cell access, creating a spatially immune-excluded phenotype (11). This immune-resistant alteration is not merely microscopic phenomena; it leaves discernable signatures on routine pretreatment computed tomography (CT) scans. Radiologic features such as tumor heterogeneity, irregular margins, and peritumoral density may serve as signs of underlying immune dysfunction (12; 13).

This convergence of tumor biology and imaging provides the conceptual foundation for radiomics. Radiomics, the extraction of quantitative features from medical images, transforms conventional imaging into numeric datasets by quantifying tumor and tissue characteristics such as shape, texture, intensity, and spatial organization (14). These features may capture aspects of the tumor microenvironment that are otherwise undetectable through conventional radiologic interpretation (15; 16). Recent studies have demonstrated the potential of utilizing radiomic signatures in clinical context. Sun et al. developed a radiomic signature that correlates with CD8+ T-cell infiltration and predicts response to anti-PD-1/PD-L1 therapy in multiple solid tumors (17). For NSCLC, radiomic features have been associated with genomic alterations, signaling pathways, and histopathology (18–25).

Our group previously reported a proof-of-concept study demonstrating that CT-derived radiomic signatures could predict both tumor response and immune-related pneumonitis in NSCLC patients treated with immune checkpoint inhibitors (ICIs), using traditional 3D features extracted with LIFEx software (26). Expanding on that concept, the present study introduces a deep learning-based radiomic framework designed to overcome the limitations of traditional feature extraction. By employing convolutional neural network (CNN)-derived layers, we aimed to capture nonlinear image patterns that may better reflect complex tumor-host interactions and immune dynamics. This approach enables an unbiased, data-driven interpretation of CT images.

This study develops and validates an artificial intelligence (AI)-based radiomic and phenomic framework to predict prognosis in advanced NSCLC treated with ICI-containing regimens. The model will target key temporal endpoints, including 12– and 36-month overall survival (OS) and progression-free survival (PFS), to support actionable risk stratification.

## METHODS

### Ethical Considerations

This investigation received institutional approval from the Northwestern University Institutional Review Board (Protocol: STU0020711, approved April 6, 2018) and adhered to principles outlined in the Declaration of Helsinki. The need for individual consent was waived due to the retrospective nature of the study.

### Patient Enrollment and Study Design

We conducted a retrospective analysis of patients diagnosed with advanced NSCLC who received ICI-containing regimen for definitive therapy at Northwestern Memorial Hospital between January 2013 and December 2022. Initial screening identified 612 potentially eligible patients.

Stringent eligibility criteria were applied to ensure radiographic suitability for quantitative analysis. Inclusion criteria were: (i) stage IV NSCLC at the initiation of treatment and (ii) measurable intrathoracic disease (≥10 mm, RECIST 1.1) suitable for radiomic characterization. Exclusion criteria included: (i) insufficient follow-up for outcome assessment, (ii) prior exposure to immunotherapy, (iii) absence of evaluable thoracic disease, (iv) suboptimal imaging quality precluding feature extraction, and (v) availability of only maximum intensity projection reconstructions. Baseline imaging and laboratory assessments followed institutional protocols, with follow-up evaluations conducted after 2-3 treatment cycles (6-8 weeks). After applying all criteria, 94 patients were included in the final analysis.

### Clinical Variable Acquisition

Demographics (age at the start of treatment; sex assigned at birth; body mass index, BMI; ECOG performance status; smoking history), pathology (histology; PD-L1 expression measured by tumor proportion score; presence of targetable driver mutations such as EGFR, ALK, ROS1 and BRAF), disease extent (number of metastatic organs; anatomical locations of metastasis), inflammatory markers (NLR, neutrophil-to-lymphocyte ratio; PLR, platelet-to-lymphocyte ratio), and treatment variables (ICI regimen; the use of concurrent platinum-based chemotherapy) were extracted from the medical records.

OS was defined as the time from initiation of the ICI-containing regimen to death from any cause, with patients censored at the date of last confirmed follow-up if alive. PFS was defined as the time from initiation of the ICI-containing regimen to radiographic progression or death, whichever occurred first. Disease progression was evaluated according to RECIST 1.1 criteria.

### Tumor Segmentation and Radiomic Feature Extraction

Baseline contrast-enhanced chest CT scans were retrieved from the institutional Picture Archiving and Communication System (PACS). All scans were acquired under standardized protocols. Manual delineation of primary tumors excluding adjacent atelectasis, pleural effusion, and normal anatomical structures was performed using LIFEx software (27) by a clinician trained in radiation oncology who was blinded to clinical outcomes.

LIFEx-derived traditional radiomic features were extracted from the finalized tumor volumes following Image Biomarker Standardization Initiative (IBSI) definitions and nomenclature. The feature set comprised (i) first-order statistics describing voxel-intensity distributions (e.g. mean, median, skewness, kurtosis, energy, entropy), (ii) shape descriptors, and (iii) texture features such as Gray Level Co-occurrence Matrix (GLCM), Gray Level Run Length Matrix (GLRLM), Gray Level Size Zone Matrix (GLSZM), and Neighboring Gray Tone Difference Matrix (NGTDM).

In addition to these LIFEx-derived radiomic features, deep learning-derived radiomic features were computed to capture multi-scale and hierarchical image patterns using a convolutional encoder-decoder architecture based on the UNET framework (28). Features were concatenated for analysis, yielding a total of 533 radiomic features.

### Feature Selection and Model Development

To address the high dimensionality of the radiomic feature space and mitigate the risk of overfitting, we implemented a rigorous feature selection and model development pipeline based on nested cross-validation. The outer loop employed five-fold cross-validation to provide an unbiased estimate of model performance, while the inner loop was used for hyperparameter tuning and feature selection.

Within each outer fold, feature selection was conducted in two sequential stages. First, univariate filtering was performed using an analysis of variance F-test to identify radiomic features significantly associated with survival outcomes, retaining those with a p-value less than 0.05. Second, multivariate selection was carried out using recursive feature elimination with cross-validation, with a Random Survival Forest algorithm serving as the base estimator. Clinical covariates were combined with the selected radiomic features to form the final feature set.

We employed an ensemble learning strategy comprising five distinct base learners: Random Survival Forest, Gradient Boosting Survival Analysis, Elastic Net penalized Cox regression, Support Vector Machine, and a deep neural network. Each learner was trained on the selected feature set, with hyperparameters optimized via Bayesian method to maximize the concordance index. Final risk predictions were generated by averaging the risk scores produced by each of the five base models.

### Model Evaluation and Statistical Analysis

The model was developed to predict 12-month and 36-month OS and PFS. Model performance was evaluated using the area under the receiver operating characteristic curve (AUROC). Additional metrics included sensitivity, specificity, positive predictive value (PPV), and negative predictive value (NPV) at optimal cutpoints determined by maximizing Youden’s index. Model calibration was assessed using calibration plots comparing predicted and observed survival probabilities. Overall predictive accuracy was quantified using the integrated Brier score (IBS) across all time points.

For clinical interpretation, patients were stratified into high– and low-risk groups based on the median cross-validated risk score. Survival distributions were compared using Kaplan-Meier analysis with log-rank tests, and hazard ratios with 95% confidence intervals (CI) were estimated using Cox proportional hazards models, adjusting for potential confounders.

Model stability was assessed through bootstrap resampling (1000 iterations) to generate 95% CIs for all performance metrics (Fig. 1). Feature importance was evaluated using permutation importance scores averaged across all folds and ensemble members. All statistical analyses were performed using Python 3.8 with scikit-learn (v1.0.2), lifelines (v0.27.0), and scikit-survival (v0.17.0) libraries, as well as R (version 4.2.0) with survival, survminer, and randomForestSRC packages for survival-specific analyses. Two-sided p-values < 0.05 were considered statistically significant.

**Figure 1.**
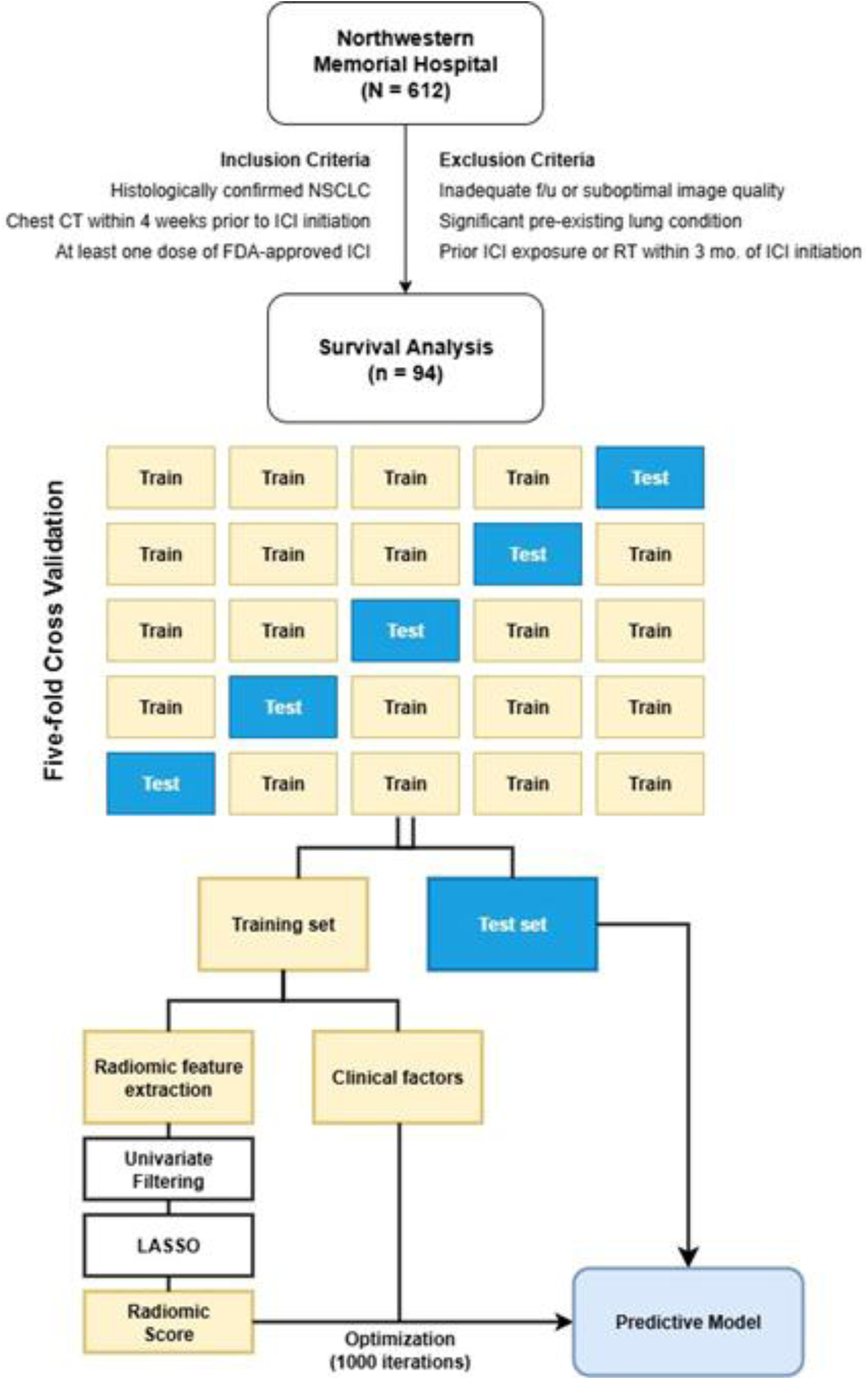
Patient Selection and Model Development Pipeline.

## Results

### Patient Characteristics

A total of 94 patients with stage IV NSCLC were included in the analysis. The median age was 66.0 years. Metastatic involvement most frequently affected the bone (n=49), pleura (n=49), contralateral lung (n=40), and brain (n=32). Pembrolizumab was the most administered ICI, followed by nivolumab, atezolizumab, and durvalumab. Approximately 40% of patients received concurrent platinum-based chemotherapy. Additional clinical variables, including PD-L1 expression and TMB, were available in subsets of the cohort. Baseline characteristics of this cohort are summarized in Table 1.

**Table 1.**
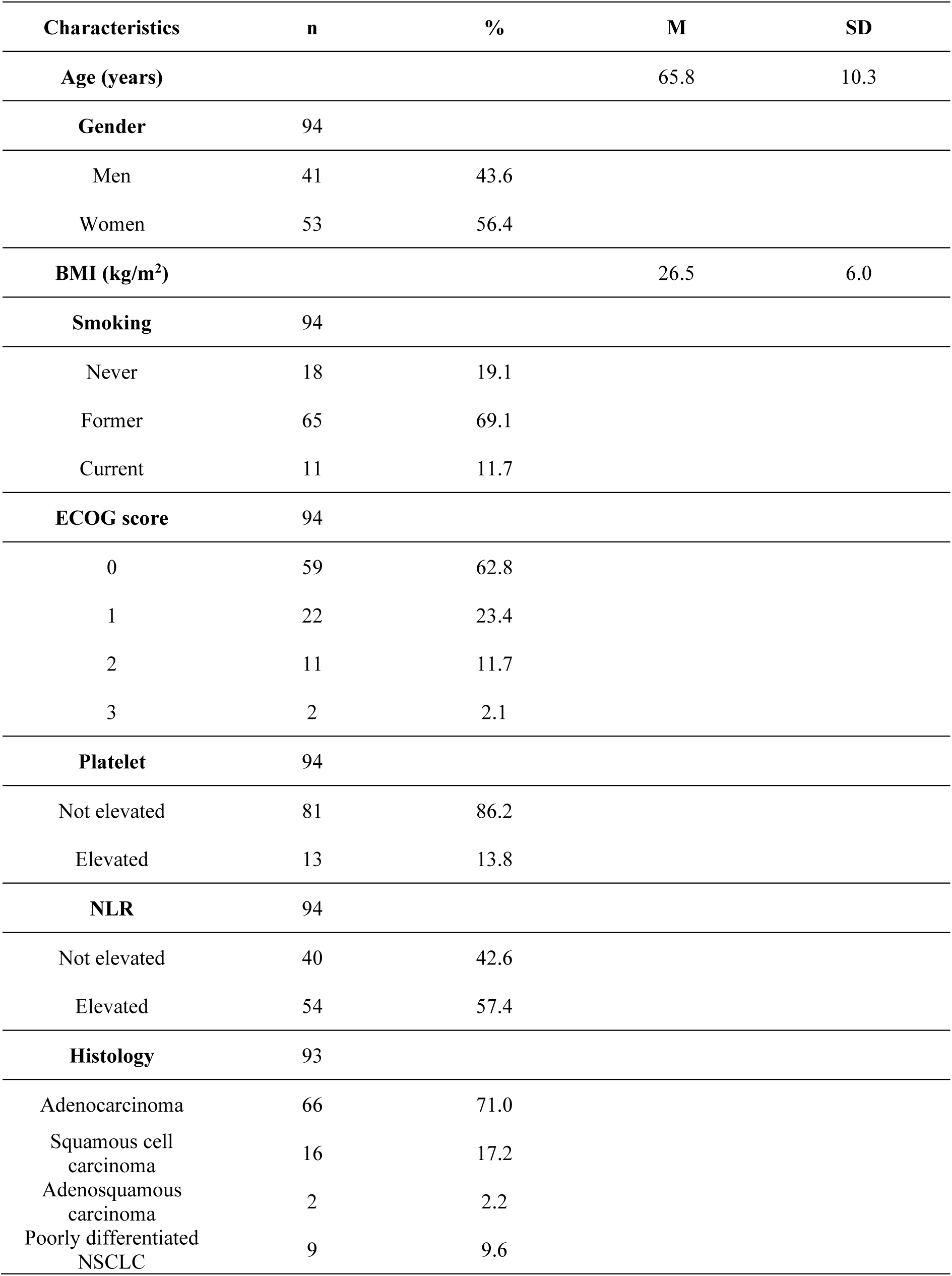
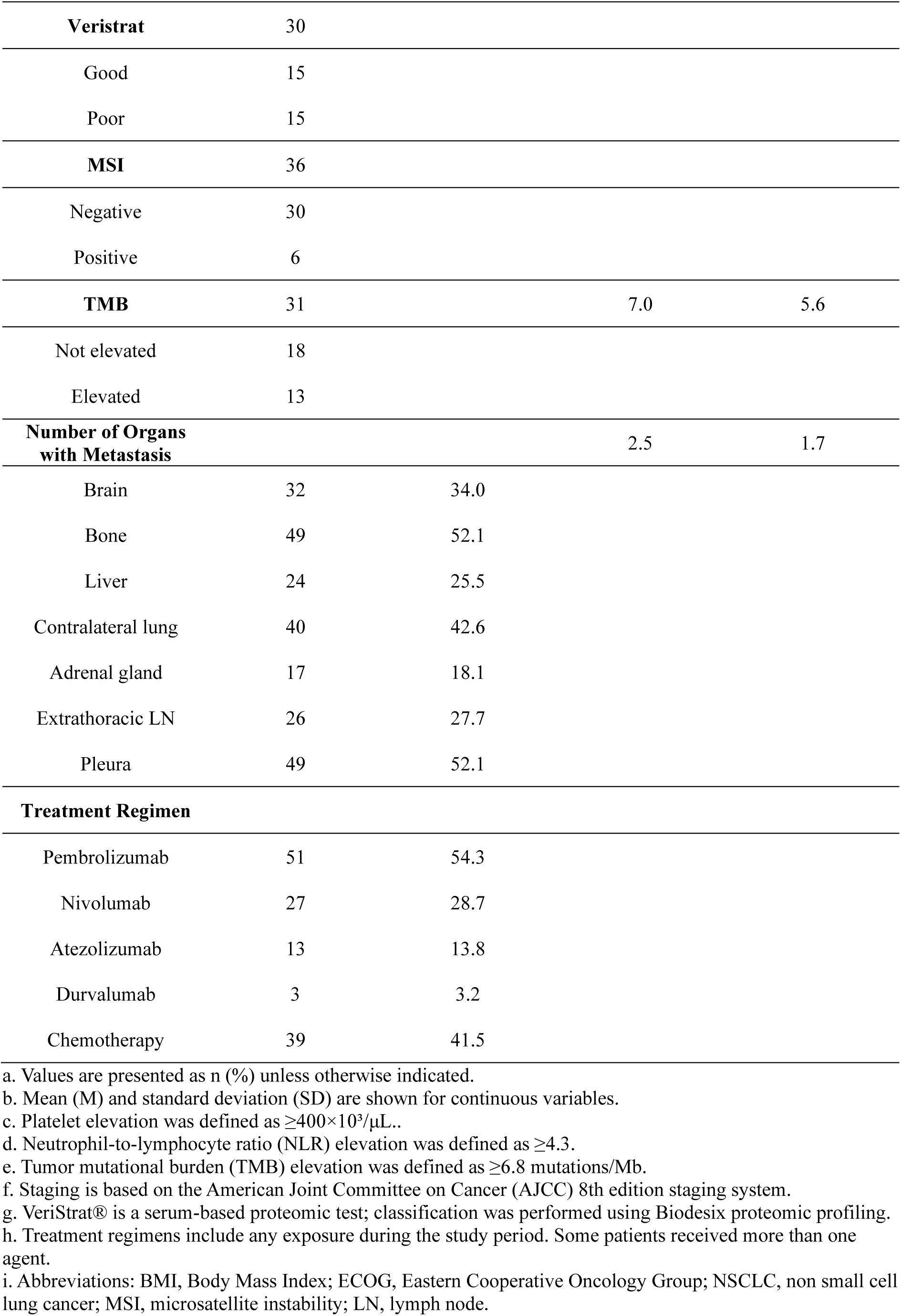
Patient Characteristics (N = 94)

### Survival Analysis

With a median follow-up of 13.5 months (IQR: 6.0–29.5), a total of 77 deaths were recorded, corresponding to a mortality rate of 81.9%. OS rates were 58.5% at 12 months and 28.7% at 36 months. Median PFS was 5.5 months, with 12– and 36-month PFS rates of 27.7% and 17.0%, respectively.

#### Model Performance

The ensemble model combining all feature categories, including clinical variables, LIFEx-based traditional radiomic features, and UNET-based deep radiomic features, demonstrated robust predictive performance for both short– and long-term survival outcomes. Figure 2 presents a representative ROC curve from one cross-validation iteration. For 12-month OS, the ensemble model achieved an average AUROC of 0.70 (95% CI: 0.65–0.75), although it is worth noting that the model integrating clinical variables with deep radiomic features reached a higher average AUROC of 0.78 during the same interval. For 36-month OS, ensemble performance improved substantially, yielding an AUROC of 0.84 (95% CI: 0.78–0.91) with Youden-optimal sensitivity and specificity of 66.0% and 90.9%. For PFS, AUROCs were 0.65 (95% CI: 0.54–0.76) and 0.73 (95% CI: 0.57–0.88) for 12– and 36-month outcomes.

**Figure 2.**
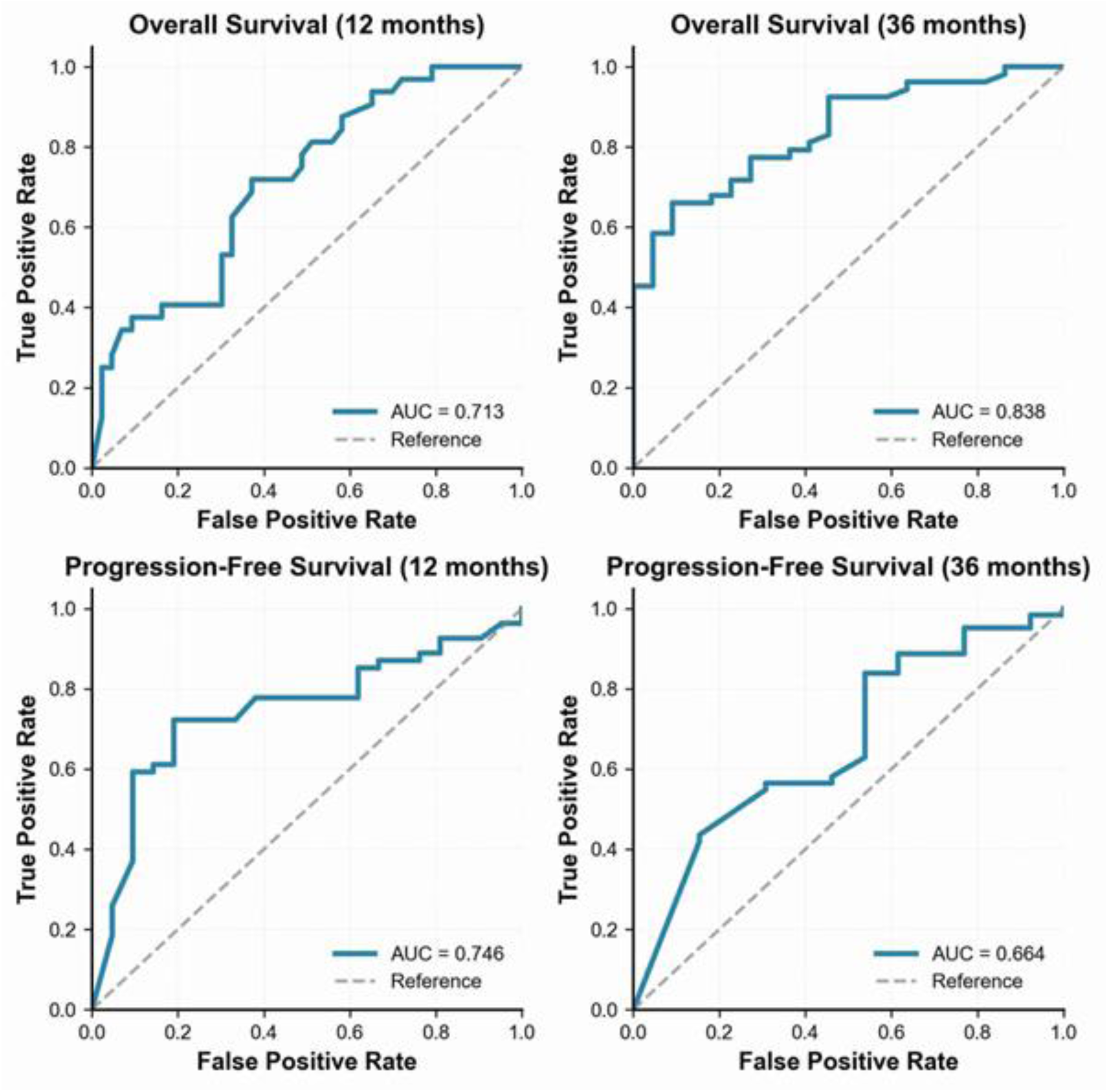
Representative ROC Curves for 12– and 36-Month Survival Generated From One Cross-Validation Iteration. Receiver operating characteristic curves for overall survival (12 and 36 months) and progression-free survival (12 and 36 months) generated by the ensemble model. Area Under Curve (AUC) values are shown for the corresponding prediction tasks.

We then compared the ensemble model with models based on clinical variables only, traditional radiomic features only, UNET-based deep features only, and their pairwise combinations (Clinical + Traditional, Clinical + UNET, and Traditional + UNET). For 12-month OS, the highest discrimination was observed in the Clinical + UNET model (Fig. 3; AUROC 0.78; 95% CI: 0.67–0.89), followed by the Traditional + UNET model (AUROC 0.76; 95% CI: 0.67–0.85) and the clinical-only model (AUROC 0.75; 95% CI: 0.67–0.83).

**Figure 3.**
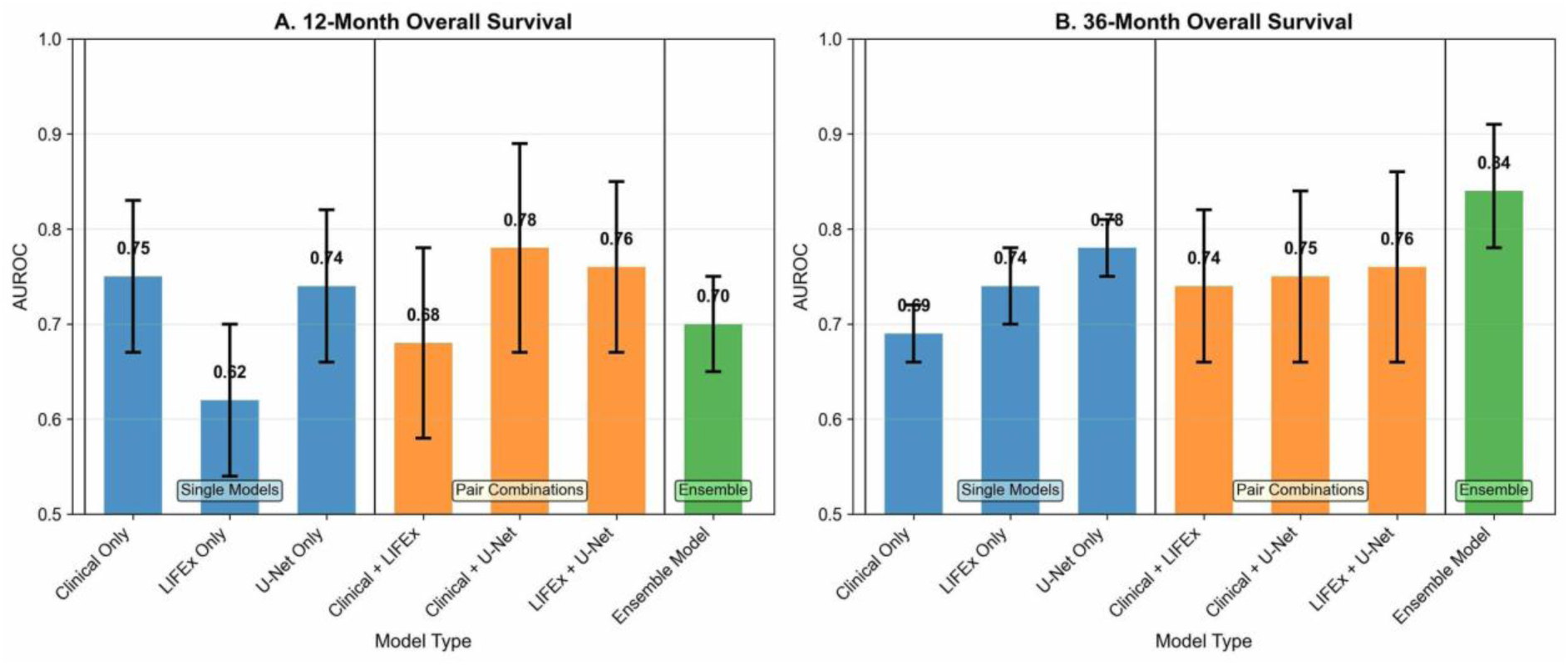
Comparison of Model Performance Using Average AUROC and 95% CI Across Feature Sets. Comparison of model performance for (A) 12-month and (B) 36-month OS using clinical variables, LIFEx-based traditional radiomics, UNET-based deep features, pairwise feature combinations, and the ensemble model. Error bars indicate 95% confidence intervals.

For 36-month OS, the ensemble model demonstrated the strongest performance (AUROC 0.84; 95% CI: 0.78–0.91). The UNET-only (AUROC 0.78; 95% CI: 0.75–0.81) and Traditional + UNET (AUROC 0.76; 95% CI: 0.66–0.86) models also showed solid discrimination, reflecting the utility of deep radiomic features in predicting long-term survival.

#### Risk Stratification

Patients were stratified into high– and low-risk groups based on the median ensemble risk score for each model. Kaplan-Meier analysis revealed significant survival differences between the two groups (Fig. 4). The median OS for the low-versus high-risk groups was 25 versus 11 months in the 12-month OS prediction model (HR 2.10; 95% CI: 1.24–3.55; p = 0.005) and 21 versus 11 months in the 36-month OS prediction model (HR 2.02; 95% CI: 1.28–3.19; p = 0.002). Similar risk discrimination was observed for PFS, with median PFS of 8 versus 3 months for both the 12-month PFS (HR 1.74; 95% CI: 1.11–2.74; p = 0.015) and 36-month PFS prediction models (HR 1.80; 95% CI: 1.15–2.82; p = 0.007).

**Figure 4.**
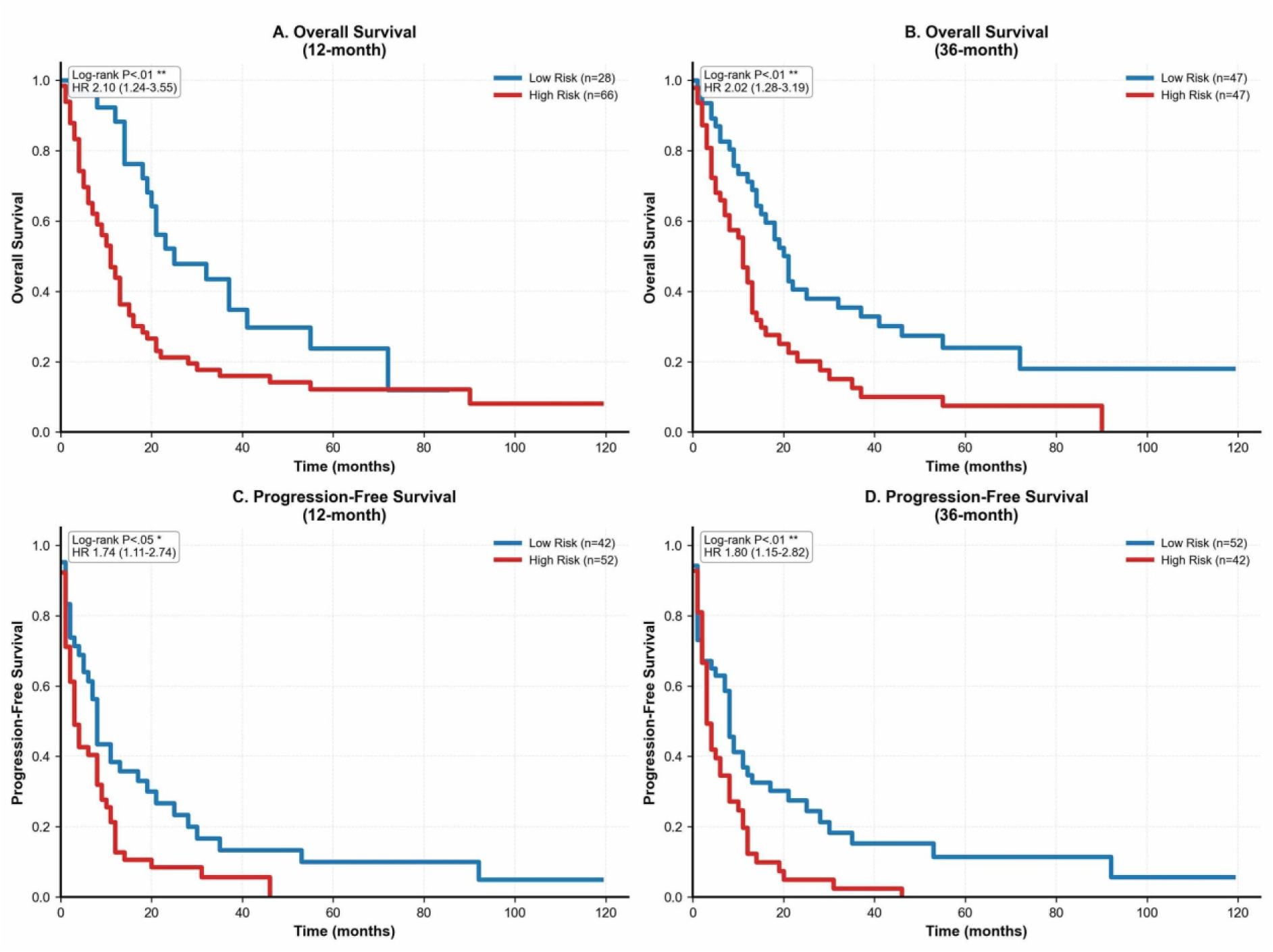
Kaplan–Meier Curves for 12– and 36-Month OS and PFS by Ensemble Model Risk Stratification. Survival analyses in low– and high-risk groups defined by the median ensemble risk score generated by the endpoint-specific model. (A) 12-month OS, (B) 36-month OS, (C) 12-month PFS, (D) 36-month PFS.

#### Feature Importance Analysis

Permutation importance analysis showed that radiomic features accounted for approximately 65% of the total model importance, whereas clinical variables contributed 35%. It is notable that while clinical factors such as BMI, NLR, and PLR were informative for predicting PFS, UNET-based and traditional radiomic features were more influential in predicting long term OS (Supplementary Fig. S1). In the 36-mo OS prediction model, the most influential feature was Deep 38 Max (importance score = 0.043), followed by NGTDM Complexity (0.038), GLCM Cluster Tendency (0.038), Intensity Histogram (0.037) and Deep 63 Std (0.035). Radiomic features reflecting tumor heterogeneity, particularly GLCM and GLSZM metrics, and measures of shape complexity, such as Morphological Compactness and GLSZM Zone Size Entropy, were informative for survival prediction. These findings imply that tumors with more irregular and heterogeneous imaging phenotypes are associated with poorer clinical outcomes.

## Discussion

This study demonstrates that a deep learning-based radiomic signature derived from baseline CT imaging can effectively predict overall and progression-free survival in patients with advanced NSCLC treated with ICI-containing regimens. Integration of radiomic features with clinical variables produced a robust predictive model, achieving AUROC value of 0.87 for 36-month survival and consistent performance across cross-validation folds.

### Biological Interpretation

It is worth noting that tumors with heterogeneous texture and irregular shape demonstrated poorer survival, indicating that radiomic metrics capture biologically meaningful aspects of immune evasion. Tumors with higher GLCM-or GLSZM-derived indices may represent microenvironmental complexity, including immunologically “cold” regions with limited T-cell access or activity. DDR1-mediated collagen remodeling, which creates extracellular barriers that exclude T-cells (29), likely underlies these imaging features. Irregular shape and loss of compactness may also indicate aggressive remodeling and epithelial-mesenchymal transition, both linked to immune escape and metastasis. Deep learning-derived features have the potential to capture additional nonlinear representations of these tumor-immune interactions not readily measurable by conventional radiomics.

### Comparison With Prior Studies

While prior clinical-only models have shown prognostic value in NSCLC immunotherapy, our approach differs in key aspects. One model reported an AUC of 0.79 for 12-month OS using both baseline and longitudinal clinical markers, whereas our model achieved comparable performance using only baseline data (30). Another study attained an AUC of 0.82 for 6-month OS based on real-world clinical variables, but performance declined at longer-term endpoints, potentially due to cohort heterogeneity and the lack of radiomic inputs (31). Diem et al. achieved an AUC of 0.74 for 10-month OS using baseline NLR and PLR but did not assess long-term OS and limited the analysis to nivolumab-treated patients (32).

Radiomics-only models also differed in focus and structure. Trebeschi et al. focused on predicting binary response to ICIs, while our model predicts OS and PFS at fixed temporal points, offering intuitive guidance for long-term care (33). Another model used baseline radiomic features and predicted 3-month OS with high sensitivity and specificity; however, it did not incorporate deep learning features or target long-term outcomes (34). Han et al.’s delta-radiomics approach improved prediction but required post-treatment imaging, limiting pre-treatment decision support (35).

A few recent studies have integrated radiomics and clinical variables like our multimodal approach (36–38). Many focused on shorter endpoints such as 12-month PFS. For example, Provenzano et al. reported an AUC of 0.79 for 24-month OS (37), while our model demonstrated higher predictive accuracy at 36 months, highlighting its strength in long-range prognostication. Together, our model shows a clear strength in delivering reliable long-range prognostication from baseline imaging alone, a meaningful advance for pre-treatment risk stratification in advanced NSCLC.

### Clinical Significance and Future Directions

The proposed model is readily translatable, relying solely on standard-of-care CT imaging and automated analysis pipelines. The semi-automatic segmentation approach, while requiring radiologist verification, could be further automated with advances in deep learning segmentation models. Feature extraction and prediction can be completed within 5–10 minutes per patient, making it compatible with clinical decision timelines. Integration with institutional imaging networks could enable real-time risk scoring at the point of care.

The potential for radiomic signature to guide adaptive treatment strategies holds important implications for future clinical practice. Radiomic risk stratification may inform treatment selection by distinguishing patients who are likely to achieve durable benefit. Patients at high risk of progression despite immunotherapy could receive closer surveillance, early combination treatment, or enrollment in clinical trials with novel agents. Such prognostic insight also helps patient goal setting, as the marked survival difference between high– and low-risk groups represents a meaningful degree of stratification that can guide care planning.

A weakness of this study is that its retrospective single-center design may limit generalizability, indicating the need for prospective, multicenter validation. Modest cohort size may have reduced power to detect covariate effects. In addition, the analysis focused exclusively on primary tumors. Future studies incorporating metastatic lesions may better capture systemic disease behavior. Although preprocessing was standardized, variations in CT acquisition and feature extraction platforms can affect reproducibility (39); harmonization across institutions will be essential for clinical translation.

Future research should focus on external validation, integration with circulating biomarkers (e.g., cell-free DNA, exosomal RNA, cytokines), and adoption of advanced architectures such as vision transformers or graph neural networks to enhance modeling of spatial dependencies and tumor-microenvironment interactions. Explainable AI techniques will improve interpretability and trust.

The path forward will require coordinated multicenter collaborations to establish standardized acquisition and analysis protocols. With protocol standardization, CT-based radiomic signatures could become a core component of precision oncology for NSCLC immunotherapy, advancing personalized treatment and patient outcomes.

## Disclosures and Acknowledgments of Research Support

J. Kim reports personal income from GeneCker Co. outside of the study. No disclosures were reported by other authors. This research did not receive any specific grants from funding agencies in the public, commercial, or not-for-profit sectors.

## Previous Presentation of the Study

The abstract for this study has been presented as a poster at the Society for Immunotherapy of Cancer (SITC) Annual Meeting 2025.

## Data Availability

All clinical data collected during the study is available upon request to the corresponding author.

