## Supplementary Figure S1 for "Integrative AI Model Combining Radiomics and Phenomics to Predict Survival in Non-Small Cell Lung Cancer Patients Treated with Immunotherapy Containing Regimen"

**Supplementary Figure S1. Feature Importance Analysis**

Permutation feature importance rankings for ensemble models. Higher values reflect greater importance.

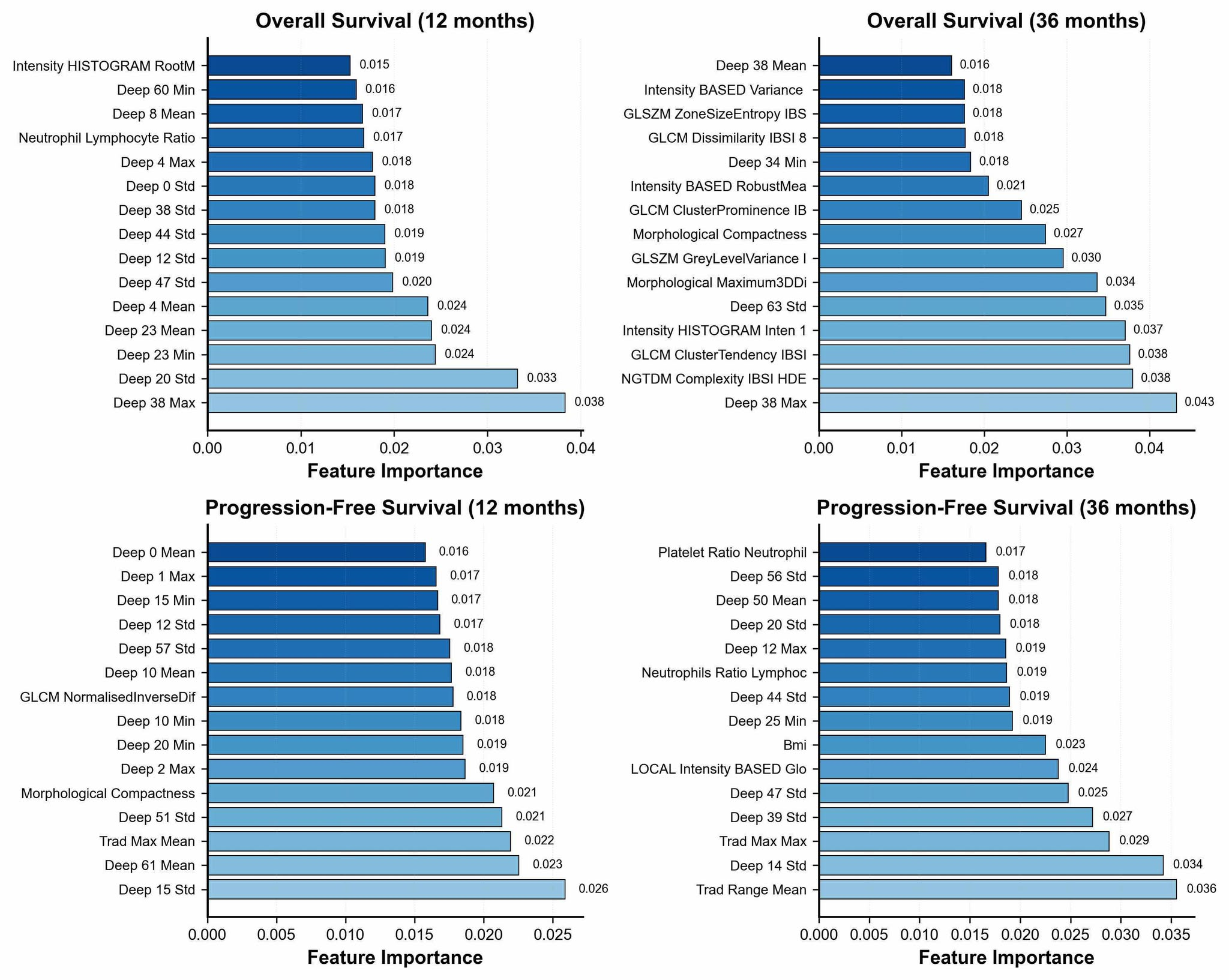
